# Risk of SARS-CoV-2 reinfection is time- and variant-dependant, France, January 2021 to August 2022

**DOI:** 10.1101/2022.11.09.22282113

**Authors:** Vincent Auvigne, Justine Schaeffer, Thibault Boudon, Cynthia Tamandjou, Julie Figoni, Isabelle Parent du Châtelet, Sibylle Bernard-Stoecklin

## Abstract

Since the emergence of Omicron, reinfections with SARS-CoV-2 have been rising. We estimated the risk of SARS-CoV-2 reinfection in the widely vaccinated French population, from January to August 2022. At nine weeks post-infection, the relative risk of reinfection, primary infection with pre-Delta variants being the reference group, was estimated at 0.43 [95%CI 0.40-0.47] if the primary infection was attributed to Delta, 0.21 [95%CI 0.19-0.24] with BA.1 and 0.17 [95% CI 0.15-0.18] with BA.2, and rapidly waned overtime. After a BA.1 primary infection the protection was similar against BA.2 or BA.4/5 reinfection.

## Introduction

Prior to the emergence of Omicron, reinfections with SARS-CoV-2 were rare [1,2]. This changed drastically during the epidemic waves driven by Omicron and its sub-lineages [2,3]. In France, only 0.7% of all COVID-19 cases detected between March and November 2021 were *possible reinfections* (hereunder *reinfections*), defined as any person with at least two positive tests performed at least 60 days apart [8,9]. In contrast, from the emergence of Omicron in December 2021 to early August 2022, this proportion increased to 6.7%, reaching 18% in week 2022-S31 [10]. Despite an increasing number of published studies on SARS-CoV-2 reinfections, a knowledge gap remains on the respective role of variant immune escape properties *versus* immunity waning, on the risk of reinfection.

This retrospective national cohort study aimed to estimate the risk of reinfection with Omicron, depending on (i) the dominant variant at the time of the primary infection, (ii) the dominant Omicron sub-lineage at the time of exposure to reinfection and (iii) the time between the primary infection and the exposure to reinfection.

### Estimating weekly risk of reinfection

Primary infections included all positive virological results (RT-qPCR, RT-LAMP, or antigen tests, excluding self-administered antigen tests) recorded in mainland France between 4 January 2021 (2021-W01) and 5 June 2022 (2022-W22). Reinfections were defined as the first positive test at least 60 days following the primary infection and between 3 January 2022 (2022-W01, beginning of the dominant circulation of Omicron in France) and 7 August 2022 (2022-W31). Individuals reinfected before this period were excluded.

Information on the variant involved in both infection and reinfections was missing for a large proportion of cases. We therefore assigned the dominant variant, defined as a variant circulating at least at 90% prevalence according to representative genomic surveillance data, [11] to each primary infection and reinfection, according to test week. For Omicron, the periods were defined for the sub-lineages BA.1, BA.2 and BA.4/5 (BA.4 and BA.5 circulated during the same period). There were no dominant variant during 12 weeks of our study period. (Figure 1).

**Figure 1:**
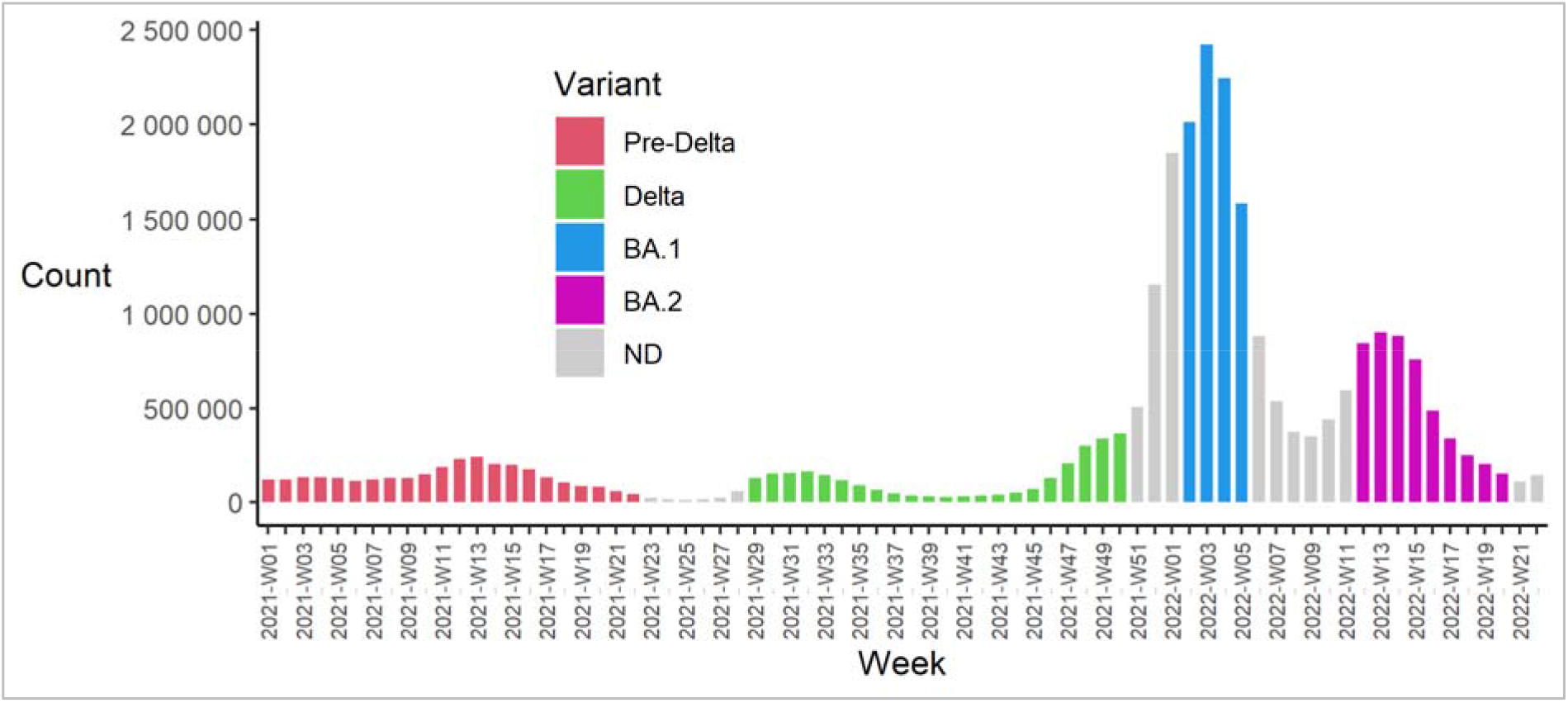
Numbers of primary infections with SARS-CoV-2 by sampling week - Mainland France - 04/01/2021 to 05/06/2022 The variants displayed are the dominant variants

We estimated the risk of reinfection for each pair of weeks, defined as the week of primary infection and the week of observation of possible reinfections (hereunder *week of exposure to reinfection*).

The steps for calculating the relative risk for each pair [i, j] are as follows. Let N_reinf_ij be the number of individuals first infected in week i and reinfected in week j and N_risk_ij the number of individuals first infected in week i who have not yet been reinfected in week j-1. The reinfection rate in week j of individuals first infected in week i is: Tx_reinf_ij = N_reinf_ij / N_risk_ij. Let Tx_reinf.Alpha_j be the reinfection rate in week j of all individuals whose primary infection occurred during the “Pre-Delta” period. Tx_reinf.Alpha_j is taken as the baseline reinfection rate for week j The relative risk of being reinfected in week j following a primary infection in week i is: HR_ij= Tx_reinf_ij / Tx_reinf.Alpha_j. This method is detailed in the supplementary.

In total, 29,652,674 individuals had at least one virological diagnosis positive for SARS-CoV-2 with a sampling date between 04/01/2021 and 05/06/2022. Among these, 71% occurred in weeks when a dominant variant could be defined. We included 3,049,290 individuals in the Pre-Delta period, 2,747,854 in the Delta period, 8,266,165 in the BA.1 period, 4,814,791 in the BA.2 period and 2,134,679 in the BA.4/5 period.

### Risk of SARS-CoV-2 reinfection is time and variant-dependant

At similar time intervals since the primary infection, relative risks differed depending on the variants involved in primary infection and reinfection (Figure 2, Table 1). Risk steadily increased from ninth week post-infection onwards, representing the time at which follow-up of reinfection started. The moving averages of relative risk following Delta and BA.1 infections were broadly parallel between 9 and 25 weeks post primary infection, indicating that the risk growth rate over time was similar for both variants. Follow-up time was not long enough to determine whether the risk of reinfection following BA.2 infection evolved in the same way. The risk of reinfection among persons first infected by BA.1 regularly increased over time, independently of Omicron sub-lineages to which those individuals were exposed. This regularity suggests that the protection conferred by a primary infection by BA.1 was similar, given the same time, against BA.2 and BA.4/5.

**Figure 2:**
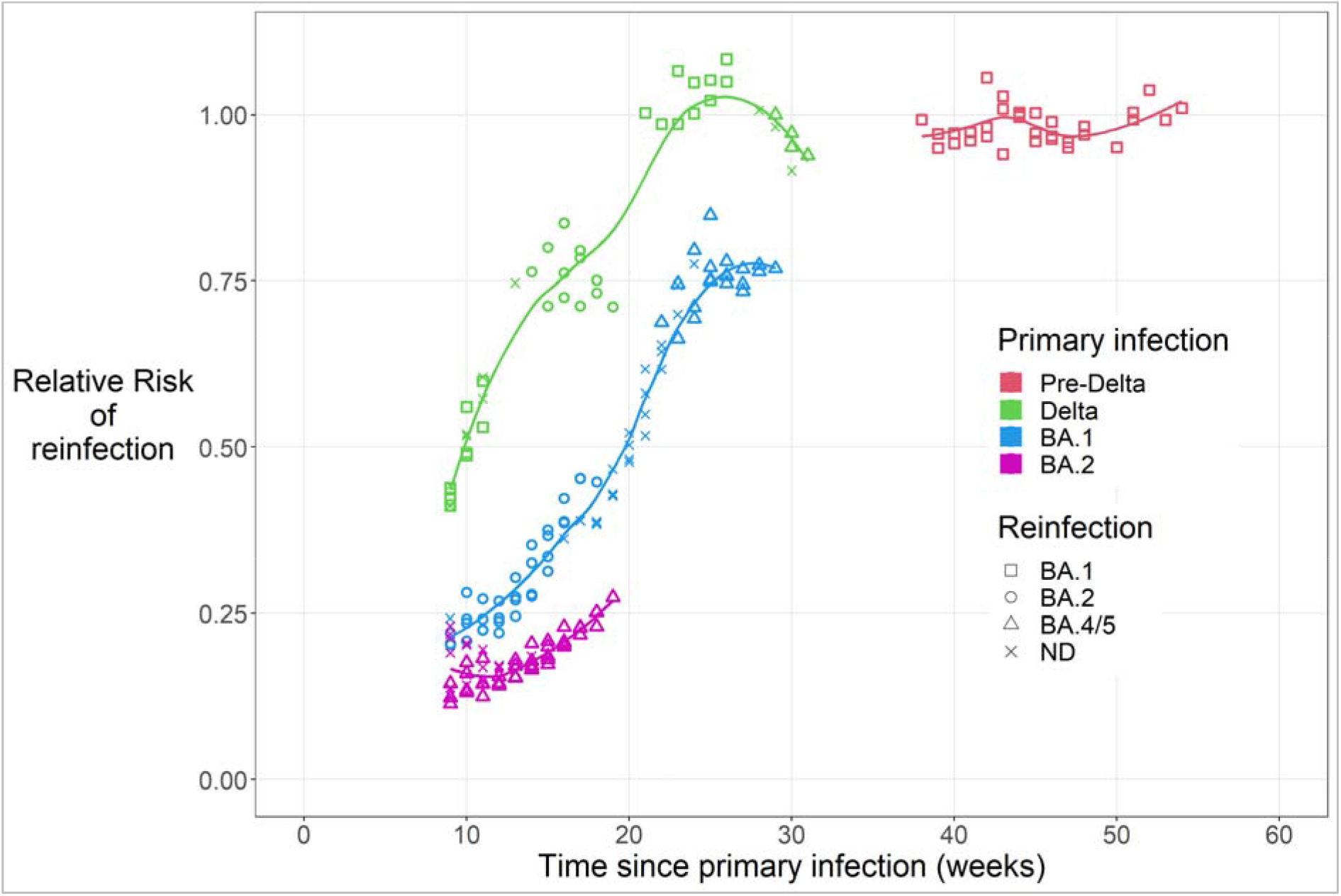
Relative Risk of reinfection by SARS-CoV-2 following a primary infection by time since primary infection, dominant variant in week of primary infection and dominant variant in week of exposure to reinfection - Mainland France - Primary infections from 04/01/2021 to 05/06/2022 - Reinfections from 03/01/2022 to 07/08/2012 Each point represents a pair of “primary infection - reinfection” weeks. The colours show the variant of the primary infection, the shapes the variant of the exposure to reinfection. [Example: 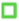 = primary infection Delta; exposure to reinfection BA.1] For each week of exposure to reinfection, the reinfection rate of individuals whose primary infection occurred during the Pre-Delta period is the reference of the Relative Risk. The pairs of weeks for which the accuracy of the estimate was low (95%CI>8%) are not represented. This parameter was set at 8% after a sensitivity study (Supplementary 2). The lines represent the moving average (Loess) for each variant of the primary infection.

**Table 1:**
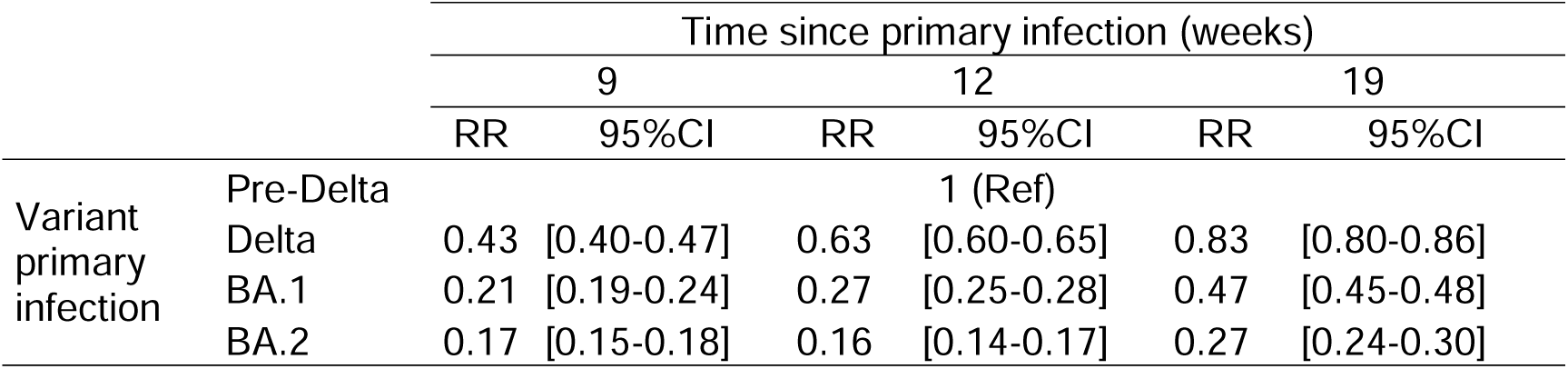
Relative Risk (RR) of reinfection following a primary infection by time since primary infection and dominant variant in week of primary infection - Mainland France - Primary infections from 04/01/2021 to 05/06/2022 - Reinfections from 03/01/2022 to 07/08/2012

|  |  | Time since primary infection (weeks) |  |  |  |  |  |
| --- | --- | --- | --- | --- | --- | --- | --- |
|  |  | 9 |  | 12 |  | 19 |  |
|  |  | RR | 95%CI | RR | 95%CI | RR | 95%CI |
| Variant primary infection | Pre-Delta |  |  | 1 (Ref) |  |  |  |
|  | Delta | 0.43 | [0.40-0.47] | 0.63 | [0.60-0.65] | 0.83 | [0.80-0.86] |
|  | BA.1 | 0.21 | [0.19-0.24] | 0.27 | [0.25-0.28] | 0.47 | [0.45-0.48] |
|  | BA.2 | 0.17 | [0.15-0.18] | 0.16 | [0.14-0.17] | 0.27 | [0.24-0.30] |

## Discussion

### Cross-protection between variants

The study shows that the risk of reinfection by Omicron and its sub-lineages depends on the variant involved in the primary infection and the variant to which the individual is later exposed. A strength of this study is the independent analysis of BA.1 and BA.2 sub-lineages. In mainland France, these two sublineages each had a distinct period of circulation while they could not be studied separately in other countries such as Denmark and Portugal [12]. We were also able to distinguish between the risk of reinfection following a primary infection by Delta *versus* BA.1. On the other hand, the sub-lineages BA.4 and BA.5 could not be distinguished from each other, as they circulated over the same period BA.5 being systematically more prevalent than BA.4. We found the lowest RR of reinfection in individuals first infected by BA.2 with a reinfection with BA.4/5 and the highest RR in those first infected with Delta and exposed to reinfection with any Omicron sub-lineage. These observations were consistent with *in silico* and *in vitro* data published elsewhere. Indeed, BA.2 and BA.4/5 are antigenically close, and it is between these sub-lineages that we observed the highest cross-protection [13]. Moreover, in agreement with our observations, the antigenic difference between Delta and Omicron is greater than between the different sub-lineages of Omicron [3]. A Portuguese national cohort study reported that the protection against reinfection with BA.4/5 is lower when the primary infection occurred in the Delta period than if it occurred in the BA.1/2 period, which is coherent with our findings. However, this study did not distinguish between the effect of time and variants [12].

### Waning of immunity over time

Our data illustrated that the immune protection against reinfection rapidly decreases over time. To our knowledge, this is the first study where the time between the primary infection and a second exposure to SARS-CoV-2 is analysed with a weekly time span. A steady increase of risk was observed from the ninth week post-infection, representing the time at which follow-up of reinfection started. This phenomenon has also been described in vaccine effectiveness studies and for post-vaccination neutralising antibodies [14–17].

### Limitations

A limitation of the study is that the vaccination status was not taken into account. On 30 May 2022 (Half of the study period), 79% of the whole French population had received at least primary series of vaccination and 59% at least one booster [18]. A study in Qatar did not observe major changes in the estimates of protection conferred by a previous infection against reinfection with Omicron before and after adjusting for vaccination status [19]. Similarly, in a prison cohort, similar protection by primary infection was observed in vaccinated and non-vaccinated individuals [20]. We therefore hypothesize that not taking into account the vaccination status of individuals does not prevent us from comparing the effect of each variant of previous infection and the time since that previous infection on the risk of reinfection. Furthermore, our method is equivalent to stratifying on the week of exposure to reinfection. In doing so, we avoid that time-dependent confounding factors, such as vaccination coverage, would bias the results. However, further studies integrating vaccination status are needed.

Virological (viral load and sequencing data) and epidemiological (contact with a confirmed COVID-19 case prior to reinfection) information required to confirm reinfections was not available. Virological tests performed in 2020 were not included; however, as reinfections were rare before the emergence of the Omicron, the likelihood that our study includes individuals for whom the first infection considered was a reinfection is low [1]. Our study did not take into account those lost to follow-up between primary infection and exposure to reinfection, which could lead to an underestimation of the risk of reinfection.

## Conclusion

We provide here real world, population-wide data on reinfections that are critical to anticipate the impact of future COVID-19 waves, and to inform public health decision makers. This approach could be relevant as a routine COVID-19 surveillance indicator. The risk of SARS-CoV-2 reinfection following a primary infection increased over time, depended on the pair of variants responsible for the first and second infections and seemed to correlate with the genetic and antigenic proximity between them. Since the beginning of 2022, the Omicron variant has been dominant, with its successive sub-lineages staying highly similar. The circulation of genetically close strains over such a long period allows good levels of cross-protection in the population. However, the emergence of a divergent variant, that would largely escape infection-induced-immunity, cannot be excluded. It is therefore important to maintain efficient genomic surveillance, allowing reactive identification of any emerging variant.

## Data Availability

While all data used in this analysis were pseudonymised, the individual-level nature of the data used risks individuals being identified, or being able to self-identify, if it is released publicly. Requests for access to the underlying source data should be directed to Sante publique France and will be granted in accordance with the GDPR and French Law.

## Ethical statement

This study did not involve the human person. It was carried out by SpF using pseudonymised data collected by the Ministry of Health to manage the COVID-19 crisis. This processing of personal pseudonymised data was implemented in accordance with the legislative and regulatory prerogatives granted to SpF to fulfil its public interest mission and in compliance with the provisions of the GDPR. In this context, the opinion of an ethics committee was not required.

## Acknowledgements

We would like to thank Benjamin Taisne, Julien Durand and Myriam Fayad for providing the data used in this study. We would also like to acknowledge all the people involved in COVID-19 surveillance activities at national and regional levels.

## Authors’ contributions

Vincent Auvigne and Sibylle Bernard-Stoecklin had full access to the data. Vincent Auvigne performed the data analysis. Vincent Auvigne, Justine Schaeffer, Thibault Boudon, Cynthia Tamandjou, Julie Figoni, Isabelle Parent du Châtelet and Sibylle Bernard-Stoecklin designed this study and contributed to data interpretation. Vincent Auvigne, Justine Schaeffer and Sibylle Bernard-Stoecklin wrote the manuscript with input from all authors. All authors made the decision to submit for publication.

