## Supplementary material for "Risk of SARS-CoV-2 reinfection is time- and variant-dependant, France, January 2021 to August 2022"

### Supplementary 1 : Calculating the Relative Risk per couple of weeks

Virological results are aggregated by 'pair of weeks'. A pair of weeks is defined by the week of primary infection (week i) and the week of exposure to reinfection (week j). The steps for calculating the relative risk for each pair [i, j] are as follows.

Let N_reinf_ij be the number of individuals first infected in week i and reinfected in week j and N_risk_ij the number of individuals first infected in week i who have not yet been reinfected in week j-1. The reinfection rate in week j of individuals first infected in week i is: Tx_reinf_ij = N_reinf_ij / N_risk_ij.

Let Tx_reinf.Alpha_j be the reinfection rate in week j of all individuals whose primary infection occurred during the "Pre-Delta" period. Tx_reinf.Alpha_j is taken as the baseline reinfection rate for week j The relative risk of being reinfected in week j following a primary infection in week i is: HR_ij= Tx_reinf_ij / Tx_reinf.Alpha_j.

The 95% confidence intervals for the relative risk is calculated. The precision of the estimate depends on the incidence rates of COVID-19 in the week of primary infection and the week of exposure to the risk of reinfection. Thus, for example, the confidence interval of the estimate is narrower for primary infections due to the Omicron variant, which caused an exceptionally intense epidemic wave, than for those due to the Delta variant. This was taken into account by retaining only those pairs of weeks for which the range of the confidence interval was below a threshold value. We set this parameter at 8% after a sensitivity study (Supplementary 2). The pairs of weeks for which the confidence interval of the relative risk is higher than 8% are therefore excluded.

### Supplementary 2 : Setting the accuracy threshold

The precision of the estimated Relative Risk of reinfection depends on the incidence rates in the week of primary infection and the week of exposure to the risk of reinfection. Thus, the confidence intervals of the estimate are narrower for primary infections due to the Omicron variant, which caused an exceptionally intense epidemic wave, than for those due to the Delta variant. We have taken this into account by plotting in Figure 1 of the main paper only those pairs of weeks for which the range of the confidence interval was below a threshold value.

A sensitivity analysis was carried out to determine the optimum for this parameter. Five values were tested, from 5% to 25%. (Figure S1 to Figure S5). It appears that the estimate of the risk of reinfection following a primary infection by the BA.1 and BA.2 sub-lineages is independent of the precision parameter. The estimate is also independent for primary infections by the Delta variant up to a time of 20 weeks. Beyond 20 weeks, the estimate of the risk of reinfection following an infection with Delta is sensitive to the parameter: if only the pairs of weeks with a very high precision are kept (CI<5%,Figure S5) the risk of reinfection following an infection with Delta is estimated only for a time of 9 weeks. When the precision threshold is less strict (CI<25%, Figure S1), the risk of reinfection following an infection with Delta is estimated up to 53 weeks and seems to decrease between 30 and 40 weeks. This late decrease of the relative risk is difficult to explain biologically and is likely to be related to fluctuations in the estimates according to their precision. The choice was therefore made not to retain them, setting the accuracy threshold at 8%.

Figure S1 :  Relative Risk of reinfection. Accuracy threshold: 25%

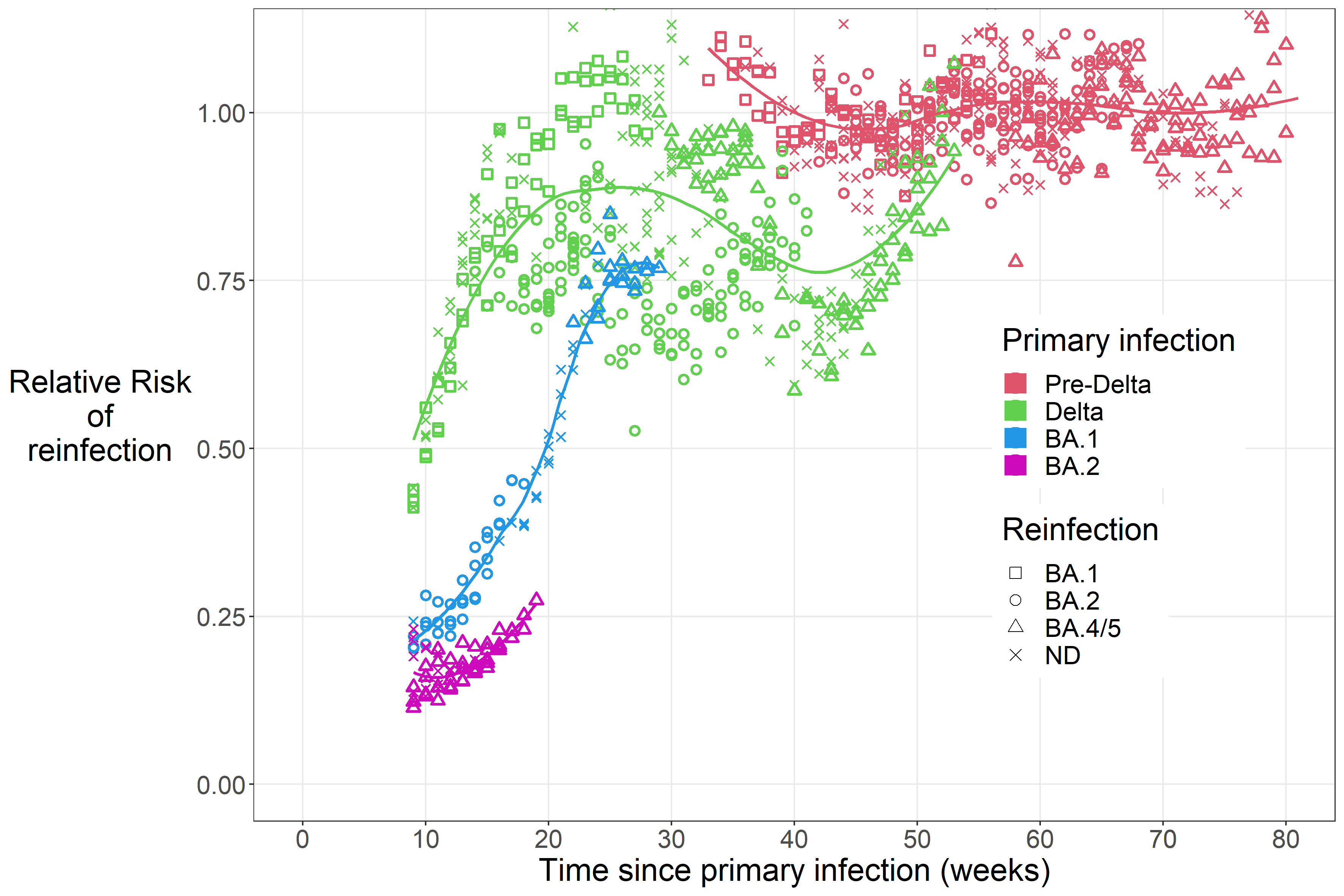

Figure S2 :  Relative Risk of reinfection. Accuracy threshold: 15%

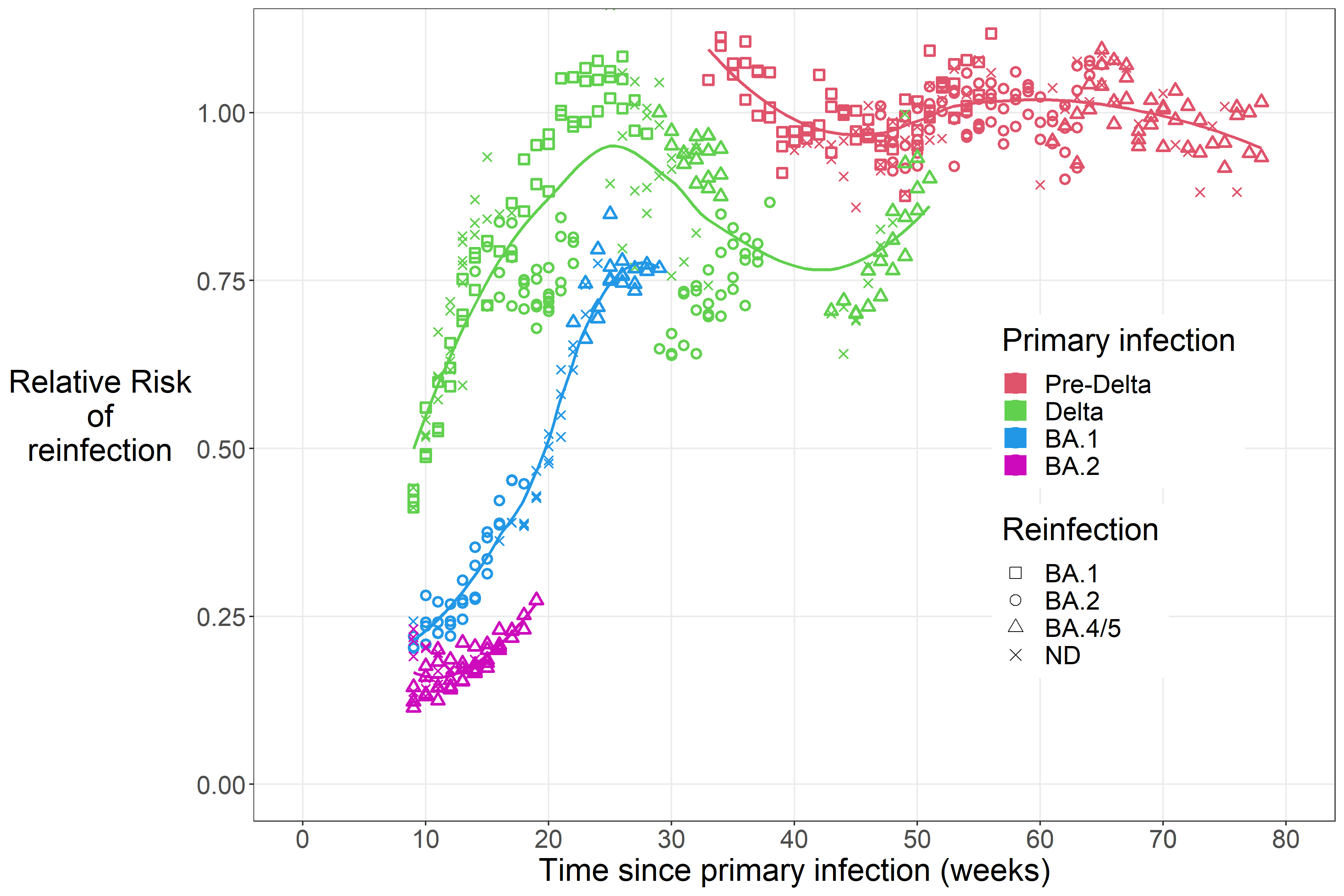

Figure S3 :  Relative Risk of reinfection. Accuracy threshold: 10%

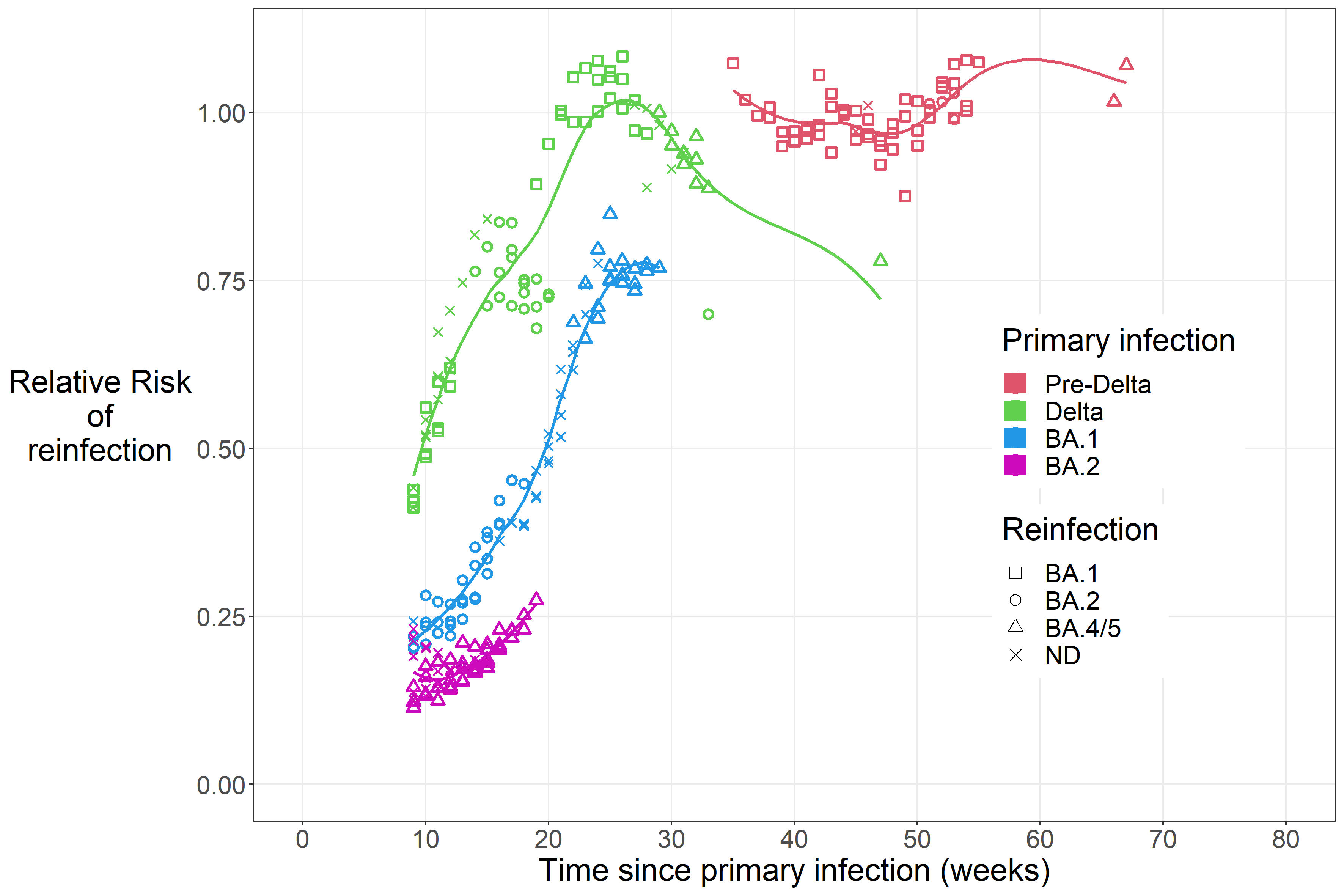

Figure S4 :  Relative Risk of reinfection. Accuracy threshold: 8%

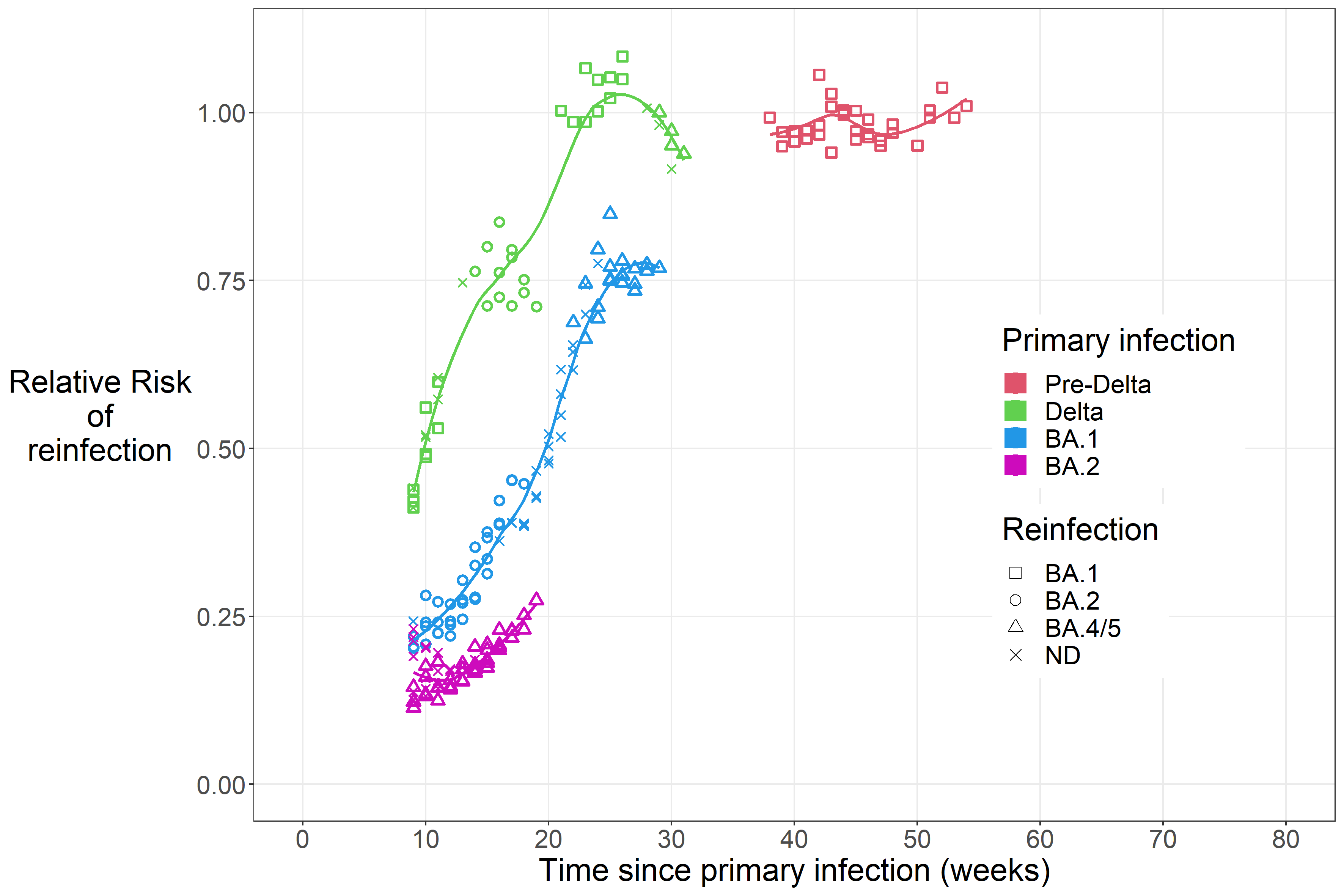

Figure S5 :  Relative Risk of reinfection. Accuracy threshold: 5%

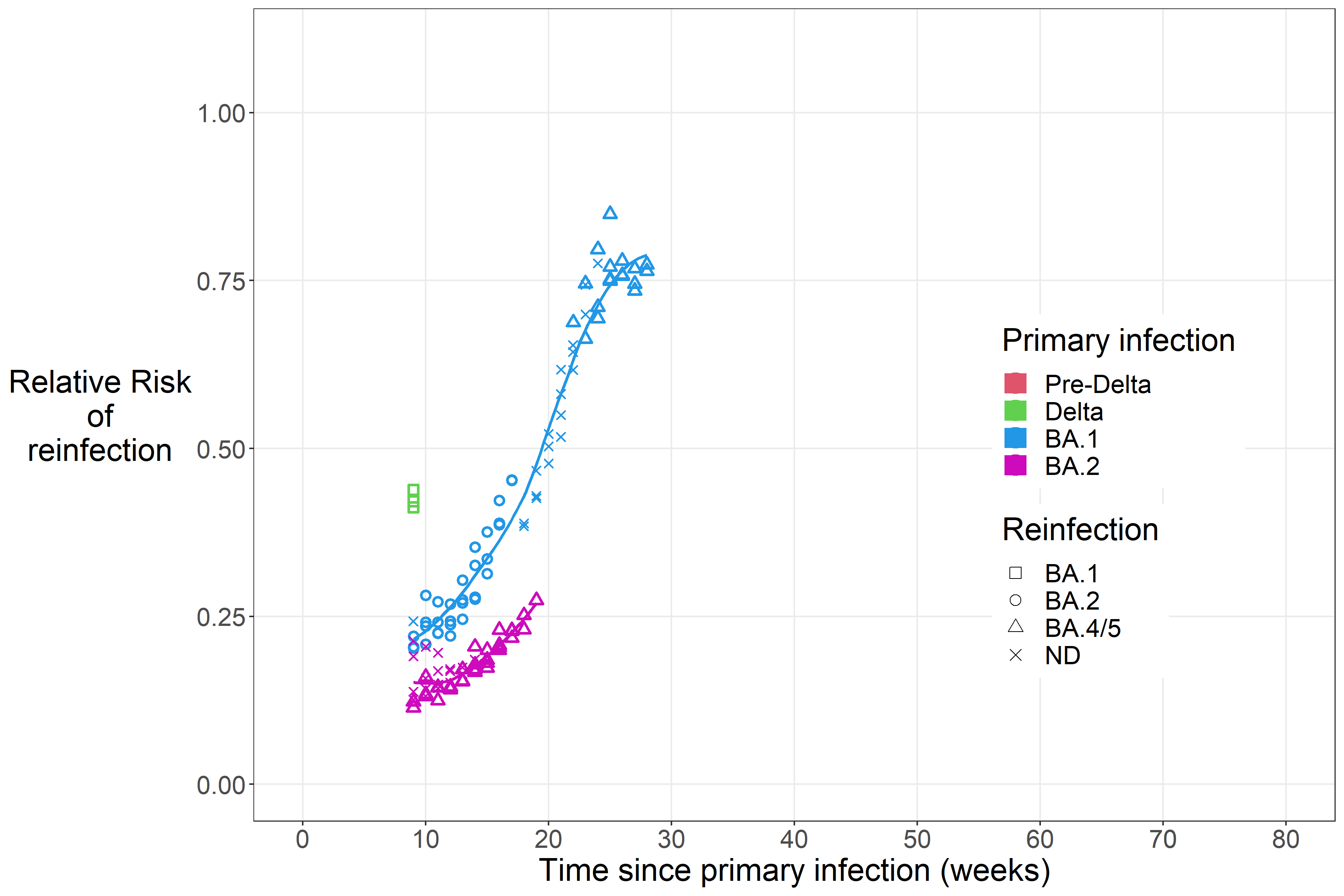
